# Attitudes and Perceptions of University Students and Postdoctoral Fellows in the Medical and Life Sciences Towards the Use of Artificial Intelligence Chatbots in the Educational Process: A Large-Scale, International Cross-Sectional Survey

**DOI:** 10.1101/2025.06.25.25328523

**Authors:** Jeremy Y. Ng, Aimun Qadeer Shah, Emi Roni, Madiha Asna, Jasleen Brar, Sylvia Kathirkamanathan, Wendy Li, Cynthia Lokker, Alfonso Iorio, R. Brian Haynes, David Moher

**Affiliations:** Centre for Journalology, Ottawa Methods Centre, Ottawa Hospital Research Institute, Ottawa, Ontario, Canada; Institute of General Practice and Interprofessional Care, University Hospital Tübingen, Tübingen, Germany; Robert Bosch Center for Integrative Medicine and Health, Bosch Health Campus, Stuttgart, Germany; Department of Health Research Methods, Evidence, and Impact, Faculty of Health Sciences, McMaster University, Hamilton, Ontario, Ontario, Canada; Department of Medicine, McMaster University, Hamilton, Ontario, Canada; School of Epidemiology, Public Health and Preventive Medicine, Faculty of Medicine, University of Ottawa, Ottawa, Canada

**Keywords:** AI chatbots, artificial intelligence, attitudes, ChatGPT, education, perceptions

## Abstract

**Background:** Artificial intelligence chatbots (AICs) are advanced systems capable of generating and processing human-like text, and are being increasingly integrated in various fields, including education. Despite their potential to significantly impact learning, little is known about university students’ and postdoctoral fellows’ (US&PD) views on AICs in educational settings. This study investigated the familiarity, perceptions, and factors influencing adoption of AICs by US&PDs in the life and medical sciences.

**Methods:** We conducted a cross-sectional online survey. Recruitment involved two approaches: (1) using R script on PubMED metadata to extract contact details of corresponding authors with recent MEDLINE-indexed publications, and (2) collecting publicly listed contact information of program administrators from the top 50 global, English-speaking universities, as ranked by the Quacquarelli Symonds (QS) list. Both authors and administrators were contacted and requested to forward the survey to US&PDs. The survey was administered via SurveyMonkey from February 2 to March 18, 2024, with two reminder emails sent between February 14 and 26, 2024.

**Results:** A total of 1,209 responses were analyzed. Most respondents identified as female (62.07%) and were enrolled in doctoral (40.48%) or master’s programs (17.55%). Over 63% were familiar with AICs, with ChatGPT being the most used (60.3%). While many recognized the educational value of AICs, concerns about reliability and integration into academia persisted. Calls for more training and institutional support were common.

**Conclusions:** The study underscores the potential and challenges of AICs in education. While enthusiasm exists, significant concerns remain about their implementation, requiring targeted training and policy development.

## Background

Artificial intelligence (AI) refers to information processing systems that display human-like intelligence to achieve specific goals with some degree of autonomy^1,2,3^. AI systems can be software based, operating in the virtual realm (e.g., search engines, face recognition systems, voice assistants such as Apple’s Siri or Amazon’s Alexa), or hardware based (e.g., computer-controlled robots, autonomous cars, drones)^1,4,5^. Although intelligence is challenging to define in both humans and machines, AI systems demonstrate hallmarks of intelligence, including perception, reasoning, decision making, performing tasks, and even learning from past mistakes^1,3^. AI is becoming increasingly prevalent in workplace, healthcare, and educational settings (e.g., AI-powered customer service representatives, clinical diagnostic tools, and personalized educational platforms)^6^.

Chatbots, which refer to computer programs that provide services through text or voice interaction, are becoming increasingly popular applications of AI^7^. While earlier versions of chatbots were limited to recognizing and analyzing existing patterns in text, newer chatbots such as ChatGPT and Perplexity are generative, meaning they can also create new data from existing patterns^8^. Chatbots can be used for a variety of purposes, including entertainment, mental health support, to inspire new ideas, to answer questions, and even execute writing or data analysis tasks^9,10,11^. Popular examples of chatbots include ChatGPT, Bing Chat, YouChat, Perplexity, and Google Bard^12^.

AI chatbots (AICs) may be of particular interest for university students and postdoctoral fellows (US&PDs) in higher educational settings as a tool to improve learning outcomes and productivity, although more evidence is needed on this topic. Chatbots can be embedded in the educational process by administrators and instructors at higher educational institutions to provide answers to frequently asked course questions, communicate course schedules and upcoming deadlines, or converse with US&PDs as part of presentations or assignment components ^13,14,15^. Chatbots can also be used informally to supplement learning. Most students in higher education have access to internet applications, making online chatbots accessible learning tools that are available outside of traditional working hours^16^. Chatbots have been shown to efficiently present complex topics to students in easy-to-understand language with the potential to support disadvantaged students or those with varied learning styles^17^. Conversations with chatbots can also be used to derive inspiration at times when students are unsure how to approach an academic task or lack ideas for coursework such as assignments and presentations^10^.

Although AI use has been implemented across many subject areas such as medicine, arts, and foreign languages, research on chatbots and their impact on learning outcomes in educational settings is limited^3,18^. A recent study examining public social media discourse on ChatGPT in education revealed that perceptions of ChatGPT are more positive than negative, although there is no consensus on whether the tool is just ‘hype’ or has future transformative potential^10^.

Despite positive views from social media users, there are some ethical and legal concerns if using AICs in higher education. Some have argued that the use of chatbots to assist with written assignments for example, is a violation of academic integrity^13,19,20^. However, most universities are still uncertain about whether the use of AI chatbots is considered cheating^21^. Generative AI has raised questions about how we define academic integrity and plagiarism^21^. Anders^22^ points out that university students can argue that using chatbots is “doing their own work” with no unfair advantage, as they applied knowledge to use a computer program that is freely available to everyone, analyzed the output, evaluated whether it met assignment requirements, and created a final product to submit. Further, as stated by Emily Hipchen, a member of Brown University’s Academic Code Committee, if plagiarism is the use of someone’s work without proper attribution, then "I don’t know that we have a person who is being stolen from”^21^.

As this technology becomes widespread, professors may dramatically shift the way they teach, such as choosing to weave ChatGPT into lesson plans^21^, or alternatively, assigning less written work to reduce concerns around chatbot use and academic dishonesty^13,20^. There are also concerns that chatbots may not report the most accurate or up-to-date information, particularly if it is a highly specialized topic^11^. There is an ongoing debate about quality standards that should be expected from chatbots (e.g., source crediting, accuracy of information), and the legal implications of using chatbots (e.g., academic dishonesty, regulations related to copyright and ownership)^11^.

We conducted an international, cross-sectional survey to better understand the current attitudes and perceptions of US&PDs in the medical and life sciences field toward the use of AICs in the educational process. The focus on this group is particularly relevant given the rapid advancements and increasing integration of AI into medical and life sciences research and clinical practice. By gaining a deeper understanding of the perceived benefits and limitations of these chatbots and factors that may affect their usage or integration by US&PDs in academic settings, the findings can help to inform policies or processes of dissemination and/or implementation of AICs in education and beyond. Given the high ethical and practical stakes involved in the use of AI in the field of medical and life sciences, insights from this group of learners are critical for shaping how AICs could be used effectively, while simultaneously addressing concerns such as misinformation and academic dishonesty. The results of this survey have the potential to inform the way that AICs are used in learning to ensure they align with the unique demands of medical and life sciences education.

## Methods

### Open Science Statement

Prior to participant recruitment, the research protocol was registered on the Open Science Framework (OSF) (https://doi.org/10.17605/OSF.IO/AQX86). We also uploaded all study materials, including a manuscript draft and raw and clean data, on OSF (https://doi.org/10.17605/OSF.IO/9M7DZ) while awaiting publication in a peer-reviewed journal.

### Study Design

We conducted an anonymous, cross-sectional open survey to investigate the attitudes and perceptions of medical and life sciences university students (i.e., undergraduate, postgraduate) and postdoctoral fellows towards the use of AICs in the educational process. All methods were performed in accordance with the Strengthening the Reporting of Observational Studies in Epidemiology (STROBE) guidelines: https://www.strobe-statement.org/.

### Sampling Framework

Two sampling strategies were used. First, corresponding authors of all articles that were indexed in the MEDLINE database in a two-month period prior to the time of searching (May 28, 2023) were contacted because we assumed that many of these authors would either be teaching courses to students or providing direct research supervision, or they might be US&PDs themselves. Authors who were a current university student (any of undergraduate, master’s, PhD) or postdoctoral fellow were encouraged to complete the survey, and faculty authors were asked to forward the survey invitation to any eligible US&PDs they were teaching or supervising. A complete list of all journals indexed in MEDLINE (approximately 5300 journals as of April 2023) was obtained, along with their NLM IDs. A search strategy of these NLM IDs was created by JYN using OVID MEDLINE and was limited to records indexed in the last two months at the time of searching. This period was chosen as researchers actively publishing between these dates are more likely to be, or have recent connections to, current US&PDs who would be eligible for this study. Authors that published any type of article were included and duplicate names were removed. AQS and ER exported the PMID numbers that were associated with the yielded records from OVID as a .csv file and inputted them into an R script created based on the easyPubMed package to capture author names, affiliated institutions, and email addresses. The “Find Full Text” function in EndNote was then used to retrieve the full text of these articles. The full text of each article was run in another R script for text recognition to extract email addresses of researchers. All results were combined into a master list and the data was cleaned to remove errors or duplicates prior to sending invitations. A template search strategy for MEDLINE is provided in **Appendix 1** of the study protocol on OSF (https://osf.io/qu5kt).

A second recruitment approach involved manually compiling publicly available contact information (name, email) of academic program administrators at the top 50 English-speaking universities globally. The top 50 schools were chosen based on the list created by Quacquarelli Symond (QS) titled “QS World University Rankings 2024: Top Global Universities”^23^. The QS list is a highly influential and trusted ranking, and assesses universities globally based on metrics such as academic reputation, employment outcomes, faculty-to-student ratio, internationalisation of student experience, and sustainability^24,25^. A pilot step was first conducted, where six authors (AQS, ER, MA, JB, SK, WL) extracted information for program administrators from the same five randomly selected schools in the top 50 QS list. The template extraction sheet used is available on OSF: https://osf.io/jm6ub. The website of each institution was first examined to identify all undergraduate, graduate, and postdoctoral programs in the field of medical and life sciences. Then, the contact information for administrators of each program were extracted from the program or faculty webpages. The extractions completed by each author were compared to ensure consistency in the program and type of contact person that was included. After the pilot step, four authors (MA, JB, SK, WL) independently and in duplicate, extracted contact information of the administrators from the remainder of the 50 top institutions in the QS list. Duplicate extractions were consolidated by a third reviewer (AQS, ER), and the program administrators at these universities were contacted to help distribute the survey to eligible US&PDs in the medical and life sciences field.

### Inclusion Criteria

Participants had to meet the following inclusion criteria to be eligible for the study: 1) currently enrolled in a university degree program (undergraduate, professional undergraduate, master’s, PhD) or currently a postdoctoral fellow and 2) studying in any area of the biomedical sciences, including, but not limited to, public health, biological sciences, dentistry, medicine, nursing, and rehabilitation therapy. Individuals not currently enrolled in a degree program or postdoctoral fellowship, or those studying outside of medical or life sciences programs, were excluded from the study.

### Participant Recruitment

As described above, we used convenience and snowball sampling to recruit US&PDs. Corresponding authors and program administrators were contacted by email in batches of 10,000. For each batch, JYN sent 1 initial email followed by 2 reminder emails, each sent 1 week apart. On February 2, 2024, the survey was released to the first batch of 10,000 contacts. The first reminder email for this batch was sent on February 14, 2024, and the final reminder on February 26, 2024. The final reminder email for the last batch of emails was sent on March 4, 2024. The survey closed two weeks after the final email was sent, providing a cool-down period from March 4 to March 18, 2024.

The emails included an invitation to participate, with a script approved by the research ethics board (REB) that detailed the study and its purpose, and a link to the survey, which was sent through email using the Mail Merge software. Recipients were encouraged to either take the study if they were eligible, or to assist by forwarding the invitation and survey link to eligible US&PDs.

### Sample Size

A response rate to the survey could not be calculated as there was no way to track how many corresponding authors of MEDLINE-indexed publications or academic program administrators forwarded the recruitment email to US&PDs. It was also not possible to determine how many US&PDs received the forwarded recruitment email from the academic program administrators and/or authors.

### Survey Instrument

The complete survey can be found in **Appendix II** of the protocol posted on OSF (https://osf.io/qu5kt). The survey was conducted using the University of Ottawa’s approved version of the SurveyMonkey software (https://www.surveymonkey.com/). The survey questions were developed by AQS and JYN based on a review of the literature and input from study authors who have experience conducting cross-sectional surveys, knowledge of educational research, and expertise in AI (CL, AI, RBH, DM). The survey was piloted by JYN, AQS, and ER prior to administration to ensure that the questions were easy to understand, and that could be completed in 15 to 20 minutes.

The survey included 30 questions, beginning with a screening question confirming eligibility. Participants that were confirmed to be eligible were then asked to sign an informed consent form before proceeding to 6 demographic-related questions: age, sex, type of academic program, country of study/work, area of study, and years of previous higher education. Then, 10 questions asked about familiarity and experience with AICs, such as whether they had used AICs for any purpose, which chatbots they used, and whether their academic institution had implemented any policies surrounding the use of AICs in academic settings. Eight questions assessed perceptions of the usefulness of AICs for educational activities such as writing papers, studying for tests, or conducting a research project. This was followed by four questions about the perceived benefits and challenges associated with AIC use in education. For example, participants were able to rate whether they agreed or disagreed with the proposed benefits of AICs, including that chatbots can help provide a personalized learning experience and real-time feedback. A final open-ended question provided an opportunity for participants to offer feedback or share any additional thoughts regarding the use of AICs in the educational process.

### Data Management and Analysis

Responses were collected using SurveyMonkey software and all data was exported and analysed using Microsoft Excel. Basic descriptive statistics such as counts and percentages were used to summarize study results. Not all respondents answered every question resulting in some missing data. Participants had the option to skip any question they preferred not to answer, and as a result, not all respondents answered every question. To address this, we analyzed and reported descriptive statistics (counts, percentages) based on the response rates for each individual question. Crosstabs tests were also conducted to determine post-hoc whether there were significant differences in attitudes and perceptions between subgroups (sex, age, academic program type, total years of higher education, primary area of study/degree major, and primary country of study).

Qualitative data from open-ended questions were analysed in duplicate by six independent authors (AQS, ER, SK, JB, WL, MA) using thematic content analysis. Discrepancies were resolved collaboratively between each pair of reviewers (AQS, ER, SK, JB, WL, MA). All coding was then reviewed by a third reviewer (AQS, ER, JYN). After reaching a consensus regarding the coding of survey responses, the codes were categorically classified and thematically grouped in an iterative process after discussions between AQS, ER and JYN.

### Ethical Considerations

The Ottawa Health Sciences Research Ethics Board (REB) approved this study (REB Number: 20230307-01H). Informed consent was obtained from all participants prior to survey completion. Participation was voluntary, any questions could be skipped at the respondent’s discretion, and participants had the right to withdraw from the study at any point prior to submitting the survey by simply closing the browser. All data was confidential and anonymous; no identifying information was explicitly collected. However, once the survey was submitted, participants were not able to withdraw or delete their responses. No monetary compensation was offered to participants for completing the survey.

## Results

Survey invitations were sent to 52,826 unique email contacts. 52,267 of these were retrieved through the MEDLINE recruitment approach and the remaining 559 were obtained from the second recruitment approach. We received 1344 submitted surveys, 1209 from participants who met the eligibility criteria. The completion rate (percentage of respondents that completed the entire survey) was 62%.

All of the raw data, where any potentially identifying information that the respondents disclosed was removed, is available on OSF (https://osf.io/bkgz7). The crosstabs are also available on OSF (https://osf.io/bse35). Demographic data of respondents are in **Table 1**. The majority of respondents were female (n=666/1073, 62%), with the majority aged 26-30 years (n=336/1075, 31.3%), followed closely by those aged 21-25 years (n=296/1075, 27.5%).

**Table 1:** Respondent Demographic Data.

| Question (Demographic) | Answer Choices (Demographic subgroup) | Number of Responses | Percent |
| --- | --- | --- | --- |
| Academic Program Type | Undergraduate degree (BSc, BA, BEng, BScN, etc.) | 128 | 11.88% |
|  | Professional undergraduate degree (MD, MBBS, DDS, PharmD, etc.) | 73 | 6.78% |
|  | Master's degree (MSc, MA, MPH, MLIS, etc.) | 189 | 17.55% |
|  | Doctoral degree (PhD, EdD, etc.) | 436 | 40.48% |
|  | MD/PhD dual degree | 30 | 2.79% |
|  | Postdoctoral fellow | 189 | 17.55% |
|  | Other (please specify) | 32 | 2.97% |
| **Total** |  | **1077** | **100%** |
| Total Years of Higher Education Completed | 0 | 16 | 1.52% |
|  | 1 | 36 | 3.42% |
|  | 2 | 44 | 4.18% |
|  | 3 | 74 | 7.03% |
|  | 4 | 111 | 10.55% |
|  | 5 | 105 | 9.98% |
|  | 6 | 114 | 10.84% |
|  | 7 | 86 | 8.17% |
|  | 8 | 82 | 7.79% |
|  | 9 | 82 | 7.79% |
|  | 10 | 89 | 8.46% |
|  | 11 | 44 | 4.18% |
|  | 12 | 52 | 4.94% |
|  | 13 | 14 | 1.33% |
|  | 14 | 19 | 1.81% |
|  | 15 | 13 | 1.24% |
|  | 16 | 20 | 1.90% |
|  | 17 | 4 | 0.38% |
|  | 18 | 11 | 1.05% |
|  | 19 | 5 | 0.48% |
|  | 20 | 7 | 0.67% |
|  | 20+ | 24 | 2.28% |
| **Total** |  | **1052** | **100%** |
| Sex | Male | 386 | 35.97 % |
|  | Female | 666 | 62.07 % |
|  | Intersex | 1 | 0.09% |
|  | Prefer not to say | 17 | 1.58% |
|  | Prefer to self-describe, please specify. | 3 | 0.28% |
| **Total** |  | **1073** | **100.00%** |
| Age | <16 | 0 | 0.00% |
|  | 16-20 | 53 | 4.93% |
|  | 21-25 | 296 | 27.53 % |
|  | 26-30 | 336 | 31.26% |
|  | 31-35 | 198 | 18.42% |
|  | 36-40 | 98 | 9.12% |
|  | 41-45 | 46 | 4.28% |
|  | 46-50 | 22 | 2.05% |
|  | 51-55 | 12 | 1.12% |
|  | 56-60 | 4 | 0.37% |
|  | 61-65 | 4 | 0.37% |
|  | >65 | 3 | 0.28% |
|  | Prefer not to say | 3 | 0.28% |
| **Total** |  | **1075** | **100%** |
| Primary Country of Study<br>(10 most common countries shown) | Brazil | 33 | 3.08% |
|  | Canada | 145 | 13.55% |
|  | China | 66 | 6.17% |
|  | India | 52 | 4.86% |
|  | Italy | 52 | 4.86% |
|  | Mexico | 35 | 3.27% |
|  | Pakistan | 45 | 4.21% |
|  | Romania | 55 | 5.14% |
|  | United Kingdom of Great Britain and Northern Ireland | 72 | 6.73% |
|  | United States of America | 200 | 18.69% |
| **\*Total** |  | **755** | **70.56%** |
| Primary Area of Study | Biology and related sciences | 490 | 45.54% |
|  | Dental studies | 47 | 4.37% |
|  | Medicine | 324 | 30.11% |
|  | Nursing | 30 | 2.79% |
|  | Medical diagnostic and treatment technology | 46 | 4.28% |
|  | Pharmacy | 43 | 4.00% |
|  | Therapy and rehabilitation | 57 | 5.30% |
|  | Other | 38 | 3.53% |
| **\*\*Total** |  | **1237** | **-** |
| *Total percent frequency does not add up to 100% as only top 20 countries shown in table. |  |  |  |
| **Because the multiple-choice question used a 'select all that apply' format, the total responses exceeded the total number of respondents. Therefore, the percent frequency of responses was not applicable. |  |  |  |

**T**he top 10 primary countries of study of respondents were led by the United States of America (n=200/946, 18.7%), Canada (n=145/946, 13.6%) and Great Britain and Northern Ireland (n= 72/946, 6.7%) (Table 1). A large portion of respondents were enrolled in a doctoral degree program (e.g., PhD, EdD) (n=436/1077, 40.5%). The most popular area of study was biology and related sciences such as biochemistry and life sciences (n=490/1076, 45.5%). The number of years of higher education that the respondents had completed was quite varied; the mode was 6 years of higher education (n=114/1052, 10.8%).

### Familiarity with the Concept of Artificial Intelligence Chatbots

As summarized in **Table 2**, most respondents indicated they were very familiar (n=216/1018, 21.2%) or familiar with the concept of AIC (n=648/1018, 63.7%), and the most popular AIC they used for any purpose was ChatGPT (n=868/1407, 85.3%). ChatGPT was also the most popular AIC used for purposes specifically relating to the educational process (n=731/1213, 72.1%).

**Table 2:** Respondent Familiarity with AI Chatbots.

| Question | Answer Choices | Frequency | Percent |
| --- | --- | --- | --- |
| How familiar are you with the concept of artificial intelligence chatbots? | Very familiar | 216 | 21.22% |
|  | Familiar | 648 | 63.65% |
|  | Unfamiliar | 100 | 9.82% |
|  | Very unfamiliar | 34 | 3.34% |
|  | I'm not sure | 20 | 1.96% |
| **Total** |  | **1018** | **100%** |
| Prior to starting this survey, which of the following AI chatbots have you used before (for any purpose)? (Please select all that apply). | Bing Chat | 161 | 15.82% |
|  | ChatGPT | 868 | 85.27% |
|  | YouChat | 23 | 2.26% |
|  | Google Bard | 155 | 15.23% |
|  | I have never used an AI chatbot | 129 | 12.67% |
|  | Other | 70 | 6.88% |
| **\*Total** |  | **1406** | **-** |
| Prior to starting this survey, which of the following AI chatbots have you used before specifically for purposes relating to the educational process (e.g., to learn concepts, to get inspiration for written work, to prepare for tests, etc.)? (Please select all that apply). | Bing Chat | 80 | 7.89% |
|  | ChatGPT | 731 | 72.09% |
|  | YouChat | 10 | 0.99% |
|  | Google Bard | 84 | 8.28% |
|  | I have never used an AI chatbot for purposes relating to scientific processes | 253 | 24.95% |
|  | Other (please specify) | 54 | 5.33% |
| **\*Total** |  | **1212** | **-** |
| *\*Because the multiple choice question used a 'select all that apply' format, the total responses exceeded the total number of respondents. Therefore, the percent frequency of responses was not applicable.* |  |  |  |

### Incorporation of Artificial Intelligence Chatbots in the Educational Process

**Table 3** summarizes the respondents views on the incorporation of AICs in the educational process, Most indicated that they were very likely (n= 377/1017, 37.1%) or likely (n=390/1017, 38.4%) to use an AIC to assist in the educational process in the future regardless of whether they had used an AIC previously.

**Table 3:**
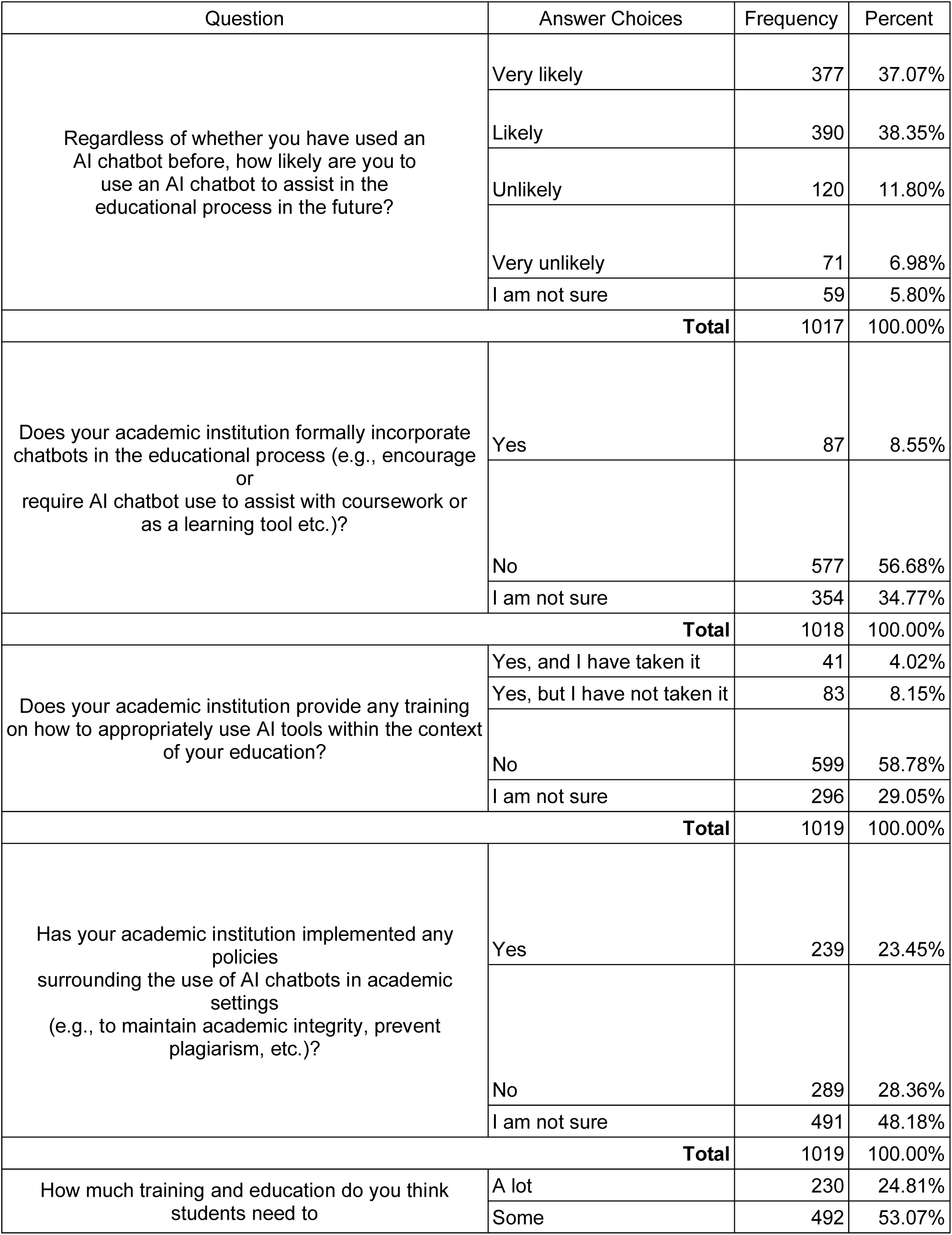
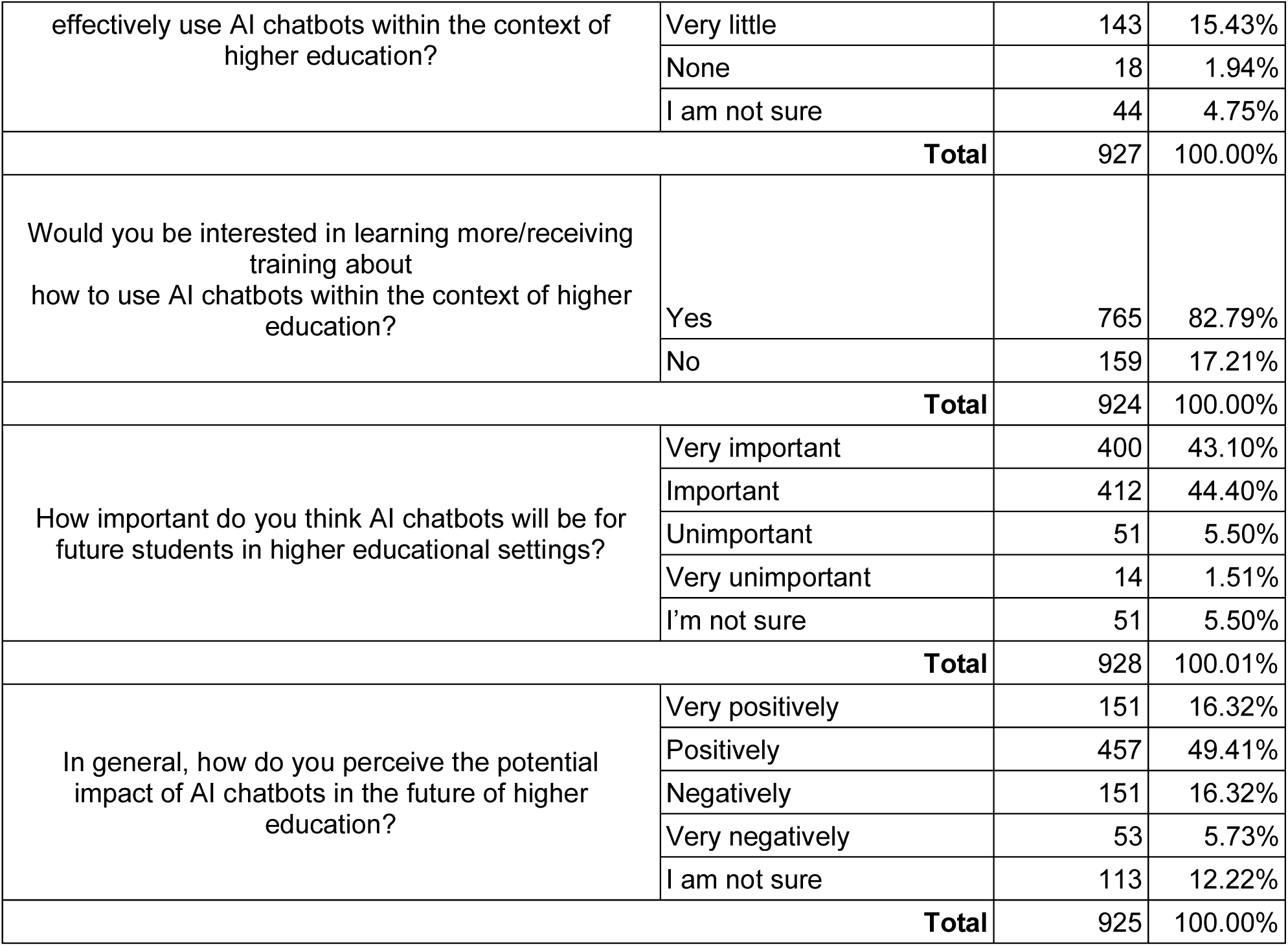
Incorporation AI Chatbots in the Educational Process.

Over half of respondents indicated that their respective academic institutions did not formally incorporate chatbots in the educational process (n=577/1018, 56.7%). They also indicated that most of their academic institutions did not provide any training on how to appropriately use AI tools within the context of education (n=599/1019, 58.8%). Many respondents were not sure of any policies about the use of AIC in academic settings implemented by their academic institutions (n=491/1019, 48.2%). About half the respondents noted that some training and education is needed for students to effectively use AI chatbots within the context of higher education (n=492/927, 53.1%), and a quarter noted that ‘a lot’ of training is needed (n=230/927, 24.8%). A large majority stated they themselves were interested in learning more/receiving training about how to use AICs within the context of higher education (n=765/924, 82.8%).

Most respondents believe AICs will be either very important (n=400/928, 43.1%) or important (n=412/928, 44.4%) for future students in higher educational settings. About two thirds positively (n=457/925, 49.4%) or very positively (n=151/925, 16.3%) perceive the potential impact of AICs in the future of higher education.

### Perceived Benefits, and Challenges of Artificial Intelligence Chatbots in the Educational Process

As seen in **Figure 1**, most respondents indicated that AICs were “helpful” or “very helpful” for the following proposed uses in the educational process: learning about a new topic (740/927, 80%), understanding a complex topic (646/925, 69.9%), and for administrative tasks such as drafting emails and creating schedules (699/926, 75.4%). On the other hand, many respondents indicated that AICs were unhelpful, very unhelpful, or that they were unsure for proposed uses such as conducting labs or experiments (727/924, 78.7%) or for conducting research or an independent study (592/926, 64%). Interestingly, when respondents were asked to indicate how often they used AICs to assist them with the same list of proposed tasks, the majority indicated that they “never,” “rarely,” or only “sometimes” used AICs for tasks such as understanding a complex topic (587/918, 63.9%) or general administrative tasks (567/920, 61.6%) (**Figure 2)**.

**Figure 1:**
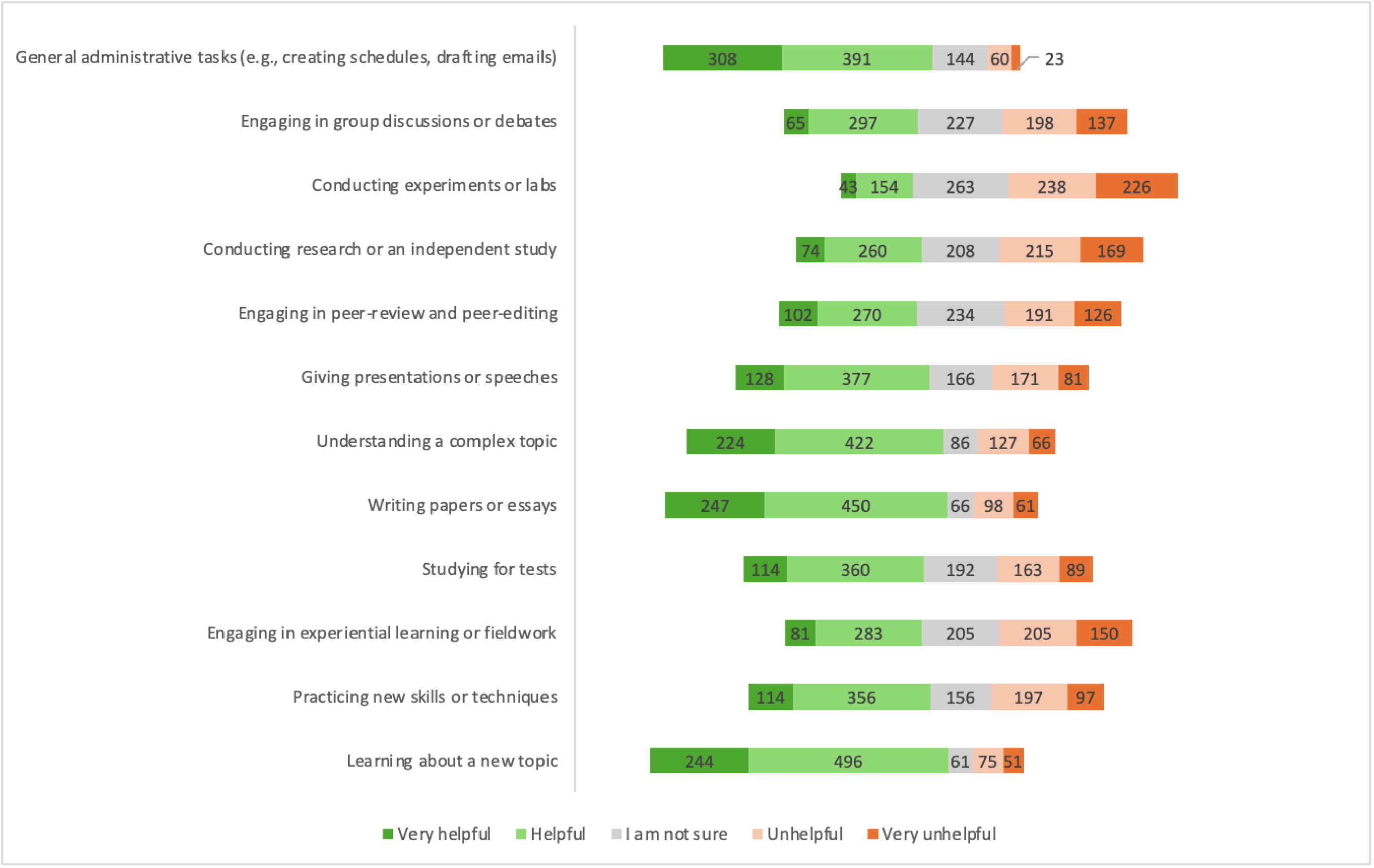
Respondent Perception of AI Chatbot Helpfulness in Different Activities that Comprise the Educational Process.

**Figure 2:**
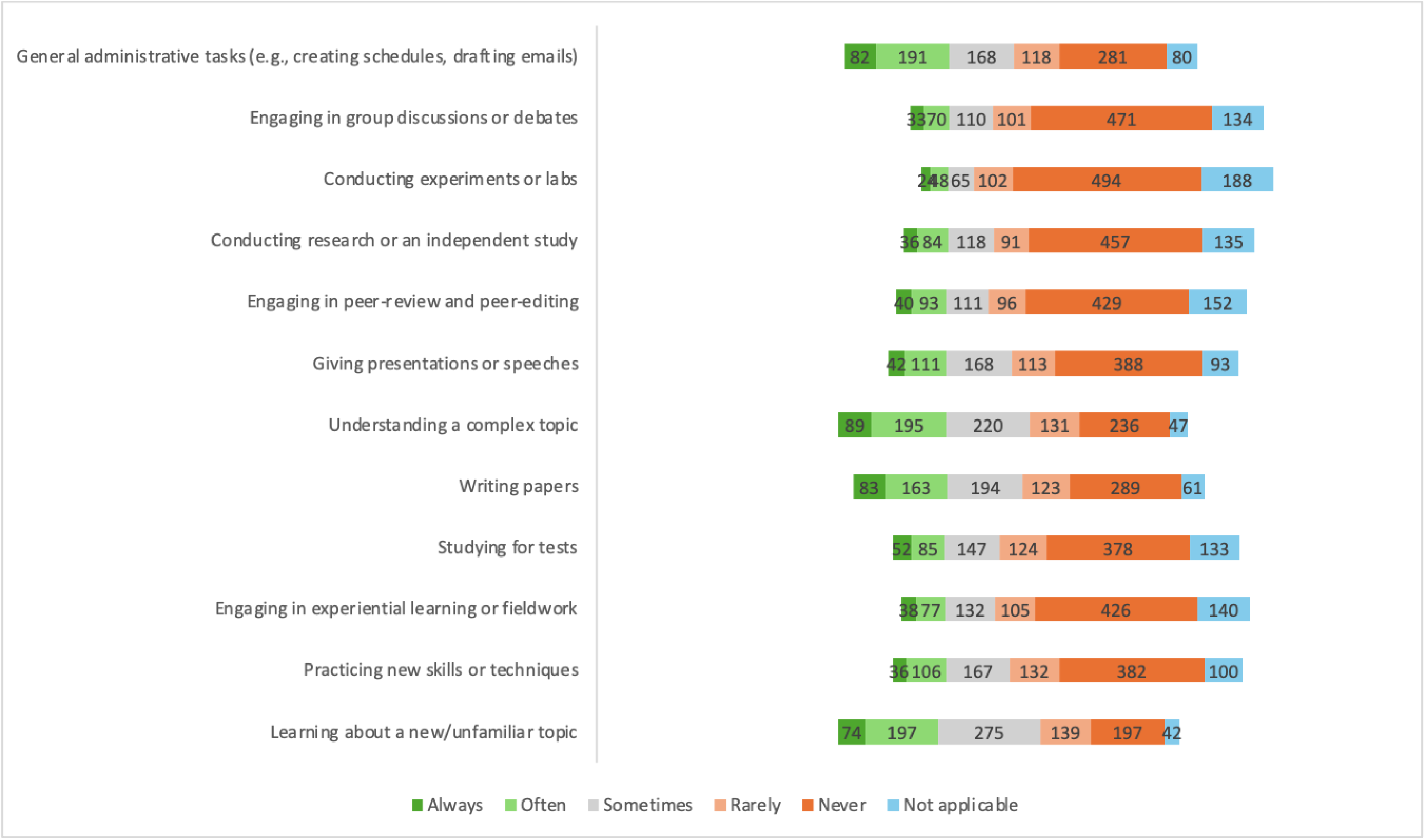
Respondent Personal Use of AI Chatbot in Different Activities that Comprise the Educational Process.

**Figure 3** shows agreement or disagreement with several listed proposed benefits of AICs in the educational process. Most notably, the majority of respondents indicated they “agree” or “strongly agree” that AICs can facilitate learning about minor subjects not directly related to the degree major, which can make it easier for students to explore new areas of interest and expand knowledge (617/825, 74.8%); that AIC use can enable 24/7 availability of educational resources and support that makes learning more convenient and accessible (648/826, 78.5%); that AICs are a cost-effective way to provide support, particularly for institutions that may have limited resources or budgets (561/826, 67.9%); and finally, that AICs can be helpful in generating ideas for creative endeavours (534/826, 64.6%). Some proposed benefits, however, did not have the same agreement. For example, most respondents indicated that they either disagreed or strongly disagreed (424/824, 51.5%) with the proposition that AICs offer consistent and reliable information to students, which can help reduce confusion about material.

**Figure 3:**
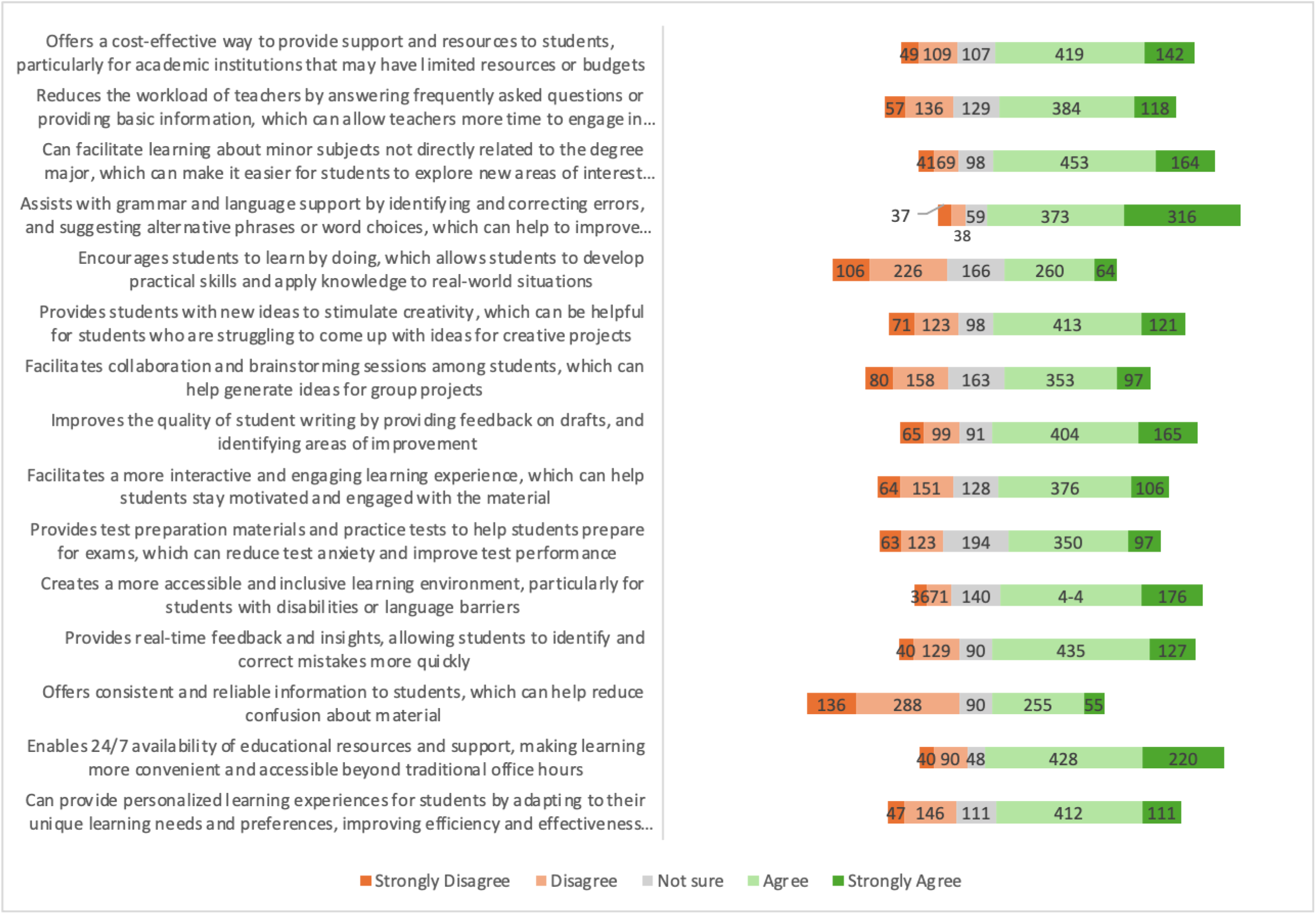
Respondent Agreement to Proposed Benefits of AI Chatbot Use in the Education Process.

As shown in **Figure 4**, most respondents indicated that they “agree” or “strongly agree” with all of the listed proposed challenges of AIC use in educational processes. For example, they indicated agreement or strong agreement that AICs can provide inaccurate or irrelevant responses due to its limitations in understanding specialised or complex subjects (671/820, 81.8%) and that school administrators have pushed against the adoption of AICs due to concerns about academic integrity (704/818, 86.1%).

**Figure 4:**
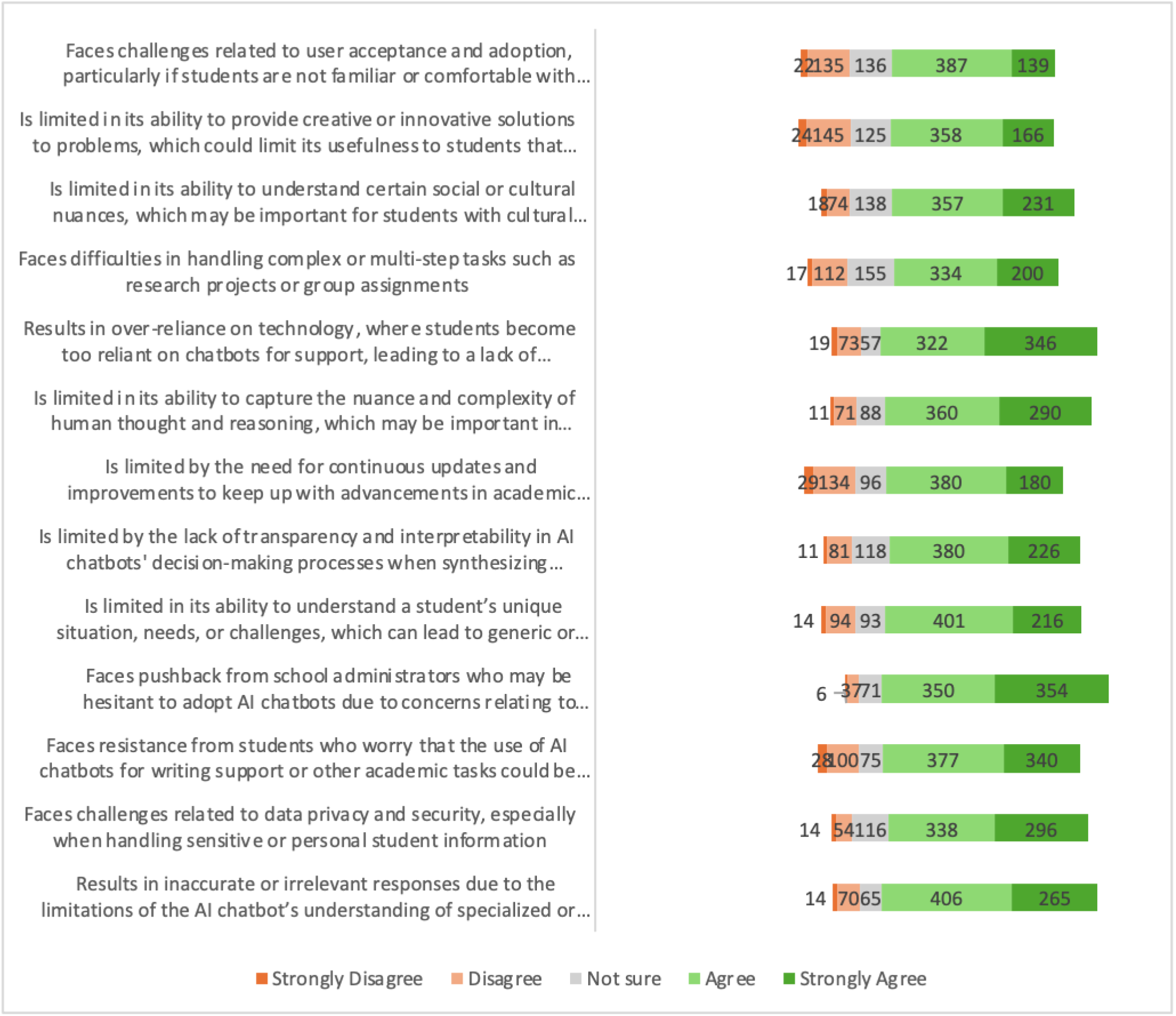
Respondent Agreement to Proposed Challenges of AI Chatbot Use in the Education Process.

### Open-Ended Questions

The open-ended questions allowed participants to provide additional comments on the following topics relating to AICs in the context of higher education: the integration and use of AICs, the nature of AIC training and its composition, institutional policies on AICs, the helpfulness of AICs, additional benefits or challenges of AICs, and final comments about the survey or the topic. **Table 4** summarizes the themes and major sub-themes identified within each subject area. Participants described the integration and use of AICs in education as supporting tasks like ‘coding,’ ‘data analysis,’ and ‘language translation,’ while raising concerns about ‘inaccuracies’ and ‘misinformation.’ Regarding institutional policies, participants discussed ‘blanket prohibitions,’ instructor-specific allowances, and the need for clearer, evolving ‘multi-level guidance.’ They highlighted the helpfulness of AICs for tasks such as ‘drafting content,’ ‘summarizing information,’ and ‘learning complex topics,’ but expressed concerns about ‘accuracy’ and ‘academic integrity.’ While some shared positive sentiments about AICs and their future potential, others offered mixed or critical reflections, emphasizing the need for caution and improved training.

**Table 4:**
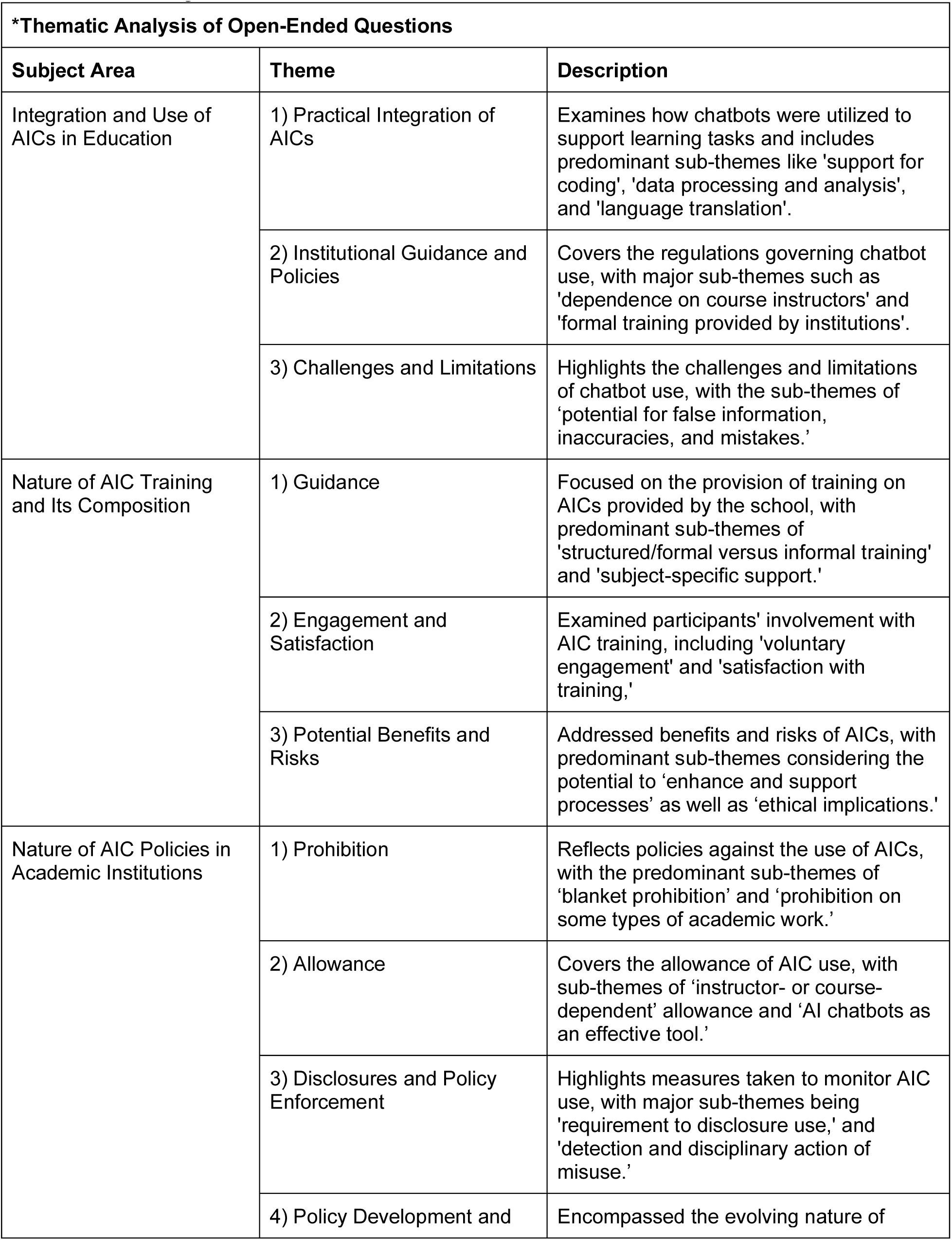
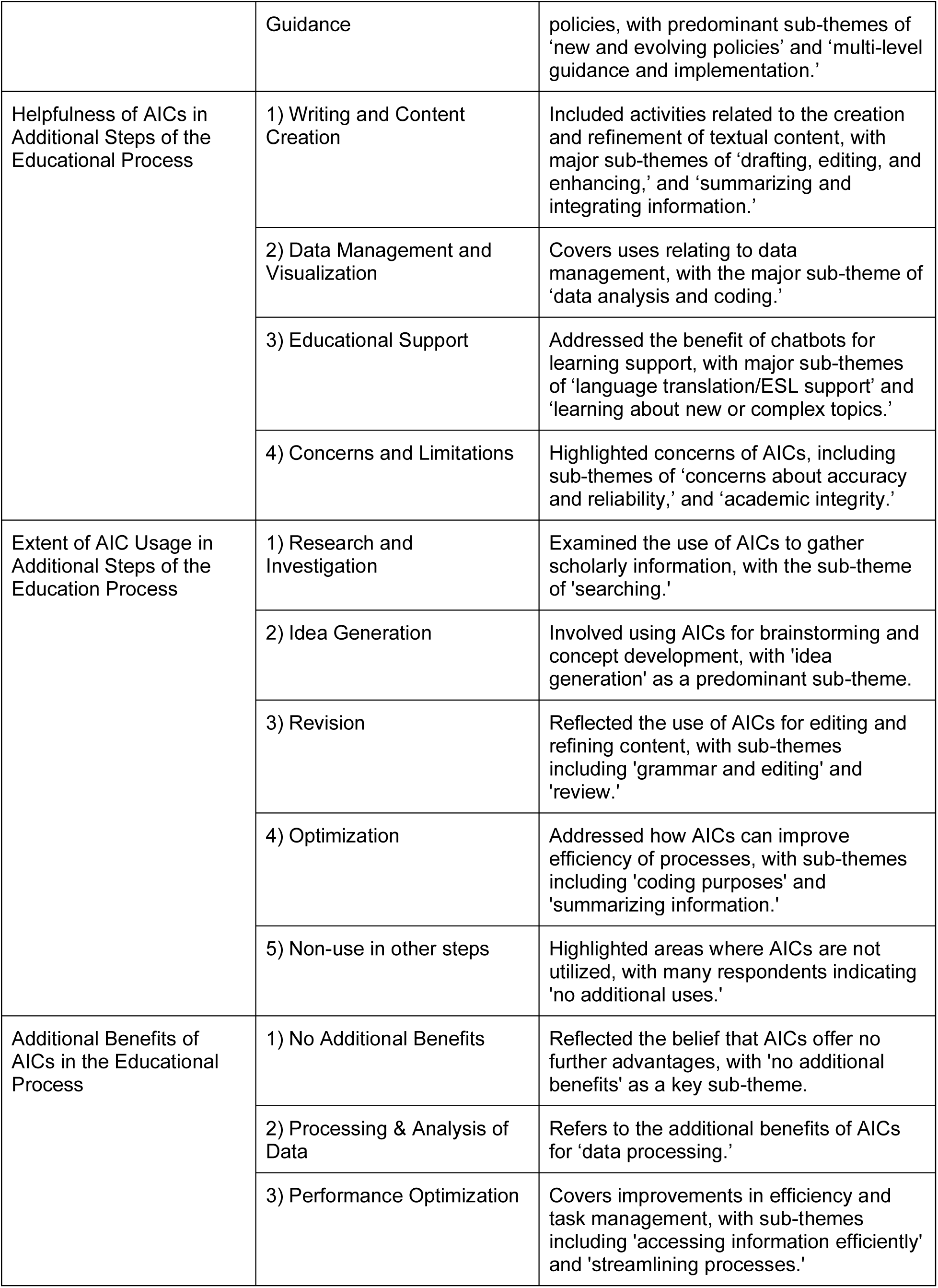
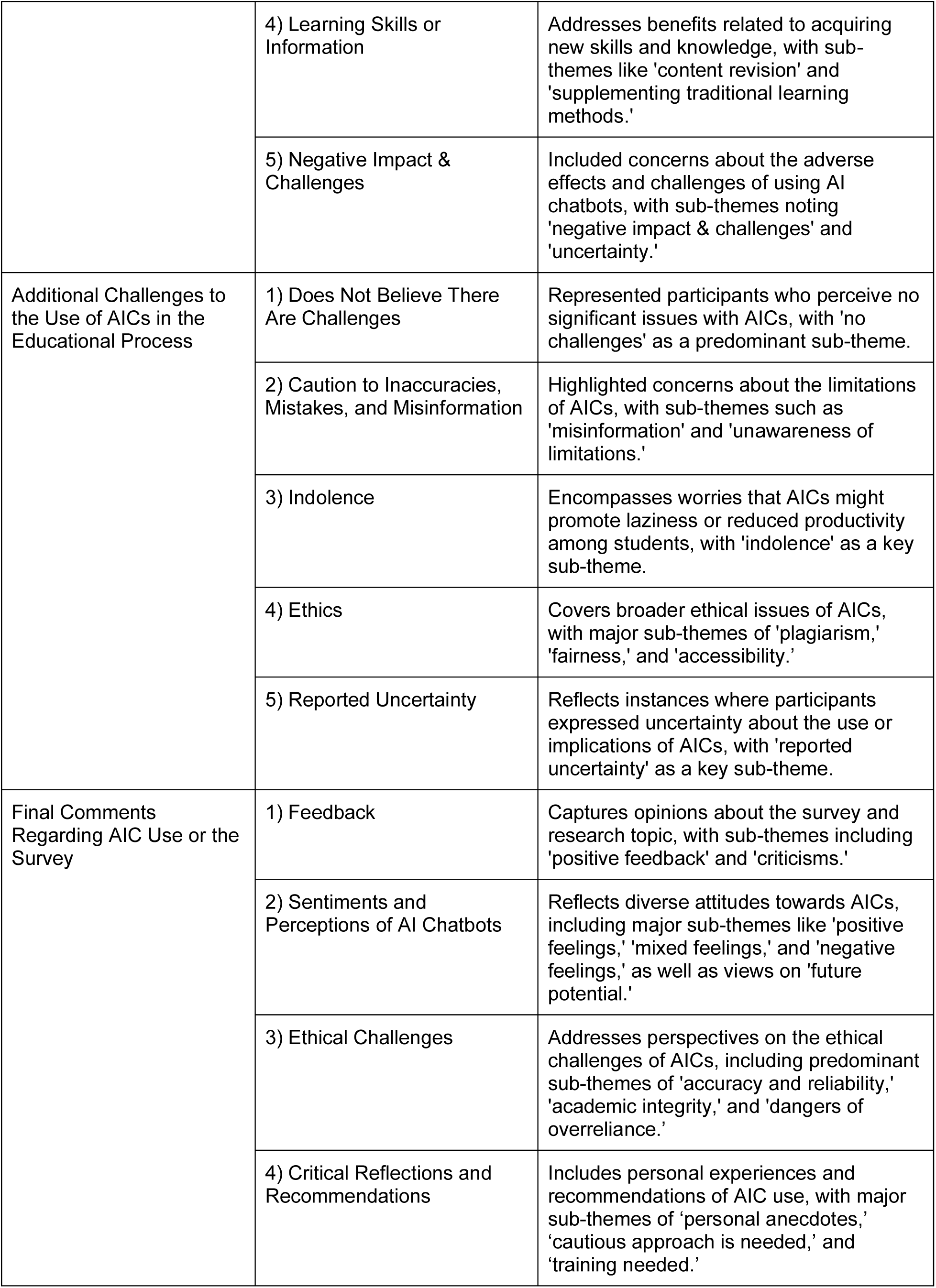
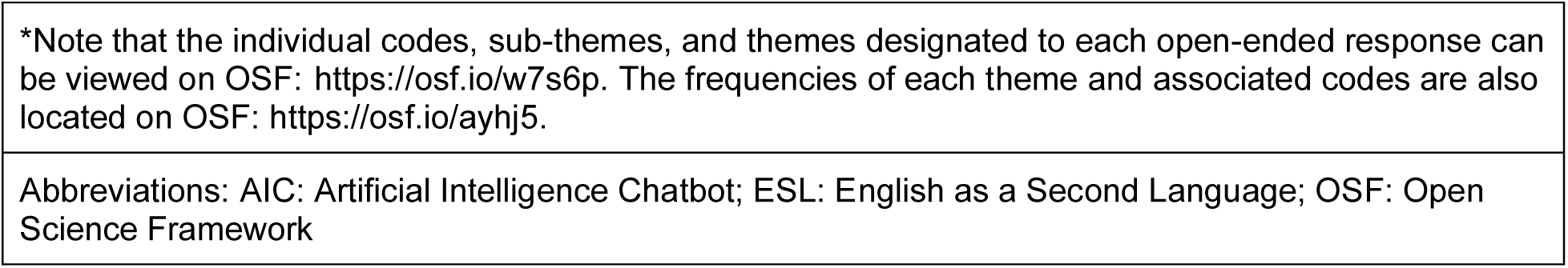
Themes and Sub-Themes from Thematic Analysis of Open-Ended Survey Questions on AI Chatbots in Education.

## Discussion

The results suggest that AICs are becoming increasingly common in the educational process. Respondents acknowledged various benefits, such as learning new or complex topics and increased accessibility to support learning beyond traditional office hours. However, they also recognized challenges, including misinformation, overreliance, inequitable access, and academic dishonesty. Additionally, respondents highlighted a range of institutional policies in response to the rise of access to AICs, from allowance of AICs with disclosure, to outright bans. Interestingly, some respondents indicated being either unaware of policies or were unable to describe them which could suggest potential gaps in communication between the institutions and learners. It may also reflect the struggle that many institutions are facing in developing and implementing policies regarding AICs for their US&PDs.

### Comparative Literature

Approximately 75% of our respondents indicated they are likely or very likely to use AICs in the educational process, which aligns with research suggesting that AI can boost motivation and engagement, enabling students to take greater agency in their own learning^26^. The perceived benefits indicated by respondents, such as improved academic performance through individualized instruction and personalized feedback, are consistent with previous studies^26,27^. Further, open-text responses revealed that some instructors encouraged students to consult AI tools as a first step. This practice allowed students to independently acquire foundational knowledge or clarify basic concepts before engaging with instructors. By leveraging AICs for these preliminary inquiries, students can come to class or office hours with more advanced questions, allowing instructors to allocate more time for lesson planning and providing detailed, individualized feedback^26,28^. In this way, AICs can potentially serve to streamline learning processes, enabling more meaningful instructor-student interactions.

One prominent issue identified in our study and corroborated by existing research is the challenge of maintaining academic integrity. The use of AIC-produced content in submitted assignments begs a greater reflection on the fine line between seeking inspiration and plagiarism. Historically, cheating might have involved copying from a website or peeking at a classmate’s work, which were more straightforward to detect and address. In contrast, the complexity and vast capabilities of AICs require a more nuanced approach to ethics and detection^29,30^. Further, while traditional examples of cheating are widely agreed upon, there is less agreement surrounding whether AI use constitutes academic dishonesty^23,23^. Our findings reflect this variation, with respondents indicating that some instructors view AI assistance as acceptable for tasks like brainstorming or editing, but not for producing graded work, while others have either formally incorporated AICs into required assignments or banned them entirely.

Accordingly, developing clear guidelines and effective training resources that establish a unified understanding of what constitutes acceptable AI use is important to reduce ambiguity and ensure fair practices across educational institutions. Respondents indicated that this was still a gap. Training programs could equip educators and students with the knowledge and skills to leverage the benefits of AICs, navigate ethical dilemmas around the use of AICs, implement effective detection methods, and maintain academic integrity in the evolving landscape of AI-enhanced education^31-33^.

### Implications

The survey results highlight the growing presence of AICs in academia and underscores the need to address the associated legal and ethical challenges. The findings suggest that AICs have the potential to significantly influence both learning outcomes and academic integrity. Institutions should consider developing and enforcing clear guidance regarding AI use to ensure it supports, rather than undermines, educational goals. This could involve creating guidelines that delineate acceptable versus unacceptable uses of AICs, thus helping to prevent misuse while promoting their benefits. At the course level, instructors can use these insights to adapt their teaching strategies. This may involve revising syllabi to incorporate AI resources, designing assessments that minimize opportunities for misuse, and offering additional support such as revision materials and extended office hours. By proactively addressing these issues, educators can foster an environment where AICs enhance the learning experience while upholding academic standards.

### Future Directions

Our findings generate several avenues for future research. First, it would be helpful to assess the range of institutional policies on AICs to gain a deeper understanding of how and why these policies were developed and implemented. Gaining a comprehensive view of current practices will inform the development of consistent recommendations for AIC use in education that optimize learning outcomes while supporting academic integrity. Additionally, insights from other academic stakeholders such as instructors and school administrators on their perceptions of AICs in the educational context will be essential to adapt and tailor future teaching practices. Finally, examining how AIC use influences student learning outcomes in the long-term will be crucial for understanding the broader impact of this innovation on educational effectiveness and student performance.

### Strengths and Limitations

A major strength of this study is that by including a large, random, and international sample of US&PDs in the medical and life sciences, our findings are more generalizable across this group. The inclusion of two distinct recruitment methods further enhanced the comprehensiveness of our study, allowing us to capture diverse perspectives from respondents across numerous institutions. Additionally, our use of the R extraction method, an established approach for semi-automatically collecting names and email addresses, minimized human error and streamlined the data collection process. The integration of both quantitative and qualitative data also allowed us to gain a deeper, more comprehensive understanding of the research question. Finally, the coding of open-ended responses was conducted independently and in duplicate, with subsequent review by a third reviewer, ensuring a rigorous and iterative thematic analysis process.

One limitation is that participants were required to be fluent in reading and writing in English due to language constraints and resource limitations of the research team. Logistic and feasibility restrictions meant that we focused on only the top 50 institutions presented in the QS ranking in the second recruitment strategy, and that medical and life sciences diploma or certificate programs were not included. We were unable to calculate an exact response rate as we lacked information on how many corresponding authors and program administrators forwarded the survey to US&PDs, and of those who did, how many US&PDs were ultimately reached. While a substantial portion of our sample included graduate students and post-doctoral fellows, there were a limited percentage of undergraduate students. An inherent limitation of cross-sectional surveys is non-response bias, which is the potential for some perspectives to be overrepresented or underrepresented in the data due to characteristics of the respondents that choose to participate compared with those that choose to not participate. For example, despite our best effort to encourage participation regardless of AI chatbot experience, individuals with strong opinions either for or against the use of AI chatbots may have been more likely to respond to the survey. This could result in underrepresentation of those with limited experience or neutral views on the subject.

Finally, the findings are subject to recall bias, where participants are limited in their accuracy to remember or report on their past experiences, such as when we asked them to recall and describe the policy and training experiences on AICs provided by their institution.

## Conclusions

This large-scale, international, cross-sectional survey provides valuable insight into the attitudes and perceptions of US&PDs in the medical and life sciences towards the use of AICs in the educational process. Our findings suggest that while there is enthusiasm about the potential for AICs to enhance learning and productivity, there are also significant concerns, particularly around academic integrity as well as the quality of information produced by these tools. The mixed views reflect the broader uncertainties and ethical considerations that accompany the rapid development and use of AICs in higher education settings. Academic institutions will need to carefully navigate these challenges, potentially rethinking traditional approaches to teaching and assessment to ensure that the use of AICs align with academic standards and values. Future research should focus on long-term impacts of AICs in the educational process and monitor the evolving perceptions of these technologies among students and educators.

## Data Availability

All study materials and data have been made available in this manuscript or on Open Science Framework.

https://doi.org/10.17605/OSF.IO/9M7DZ

## List of Abbreviations

AI: artificial intelligence
AIC: artificial intelligence chatbot
ChatGPT: chat generative pre-trained transformer
ESL: English as a second language
OSF: Open Science Framework
REB: research ethics board

## Declarations

### Ethics Approval and Consent to Participate

Ethics approval was granted by the Ottawa Health Science Network Research Ethics Board (REB Number: 20230307-01H).

### Availability of Data and Materials

All study materials and data have been made available in this manuscript or on Open Science Framework: https://doi.org/10.17605/OSF.IO/9M7DZ

### Author Contributions

JYN conceptualized the study. JYN coordinated and managed data curation. AQS, ER, MA, JB, SK, and WL participated in formal analysis of the data. JYN, AQS, and ER contributed to writing of the original manuscript draft. CL, AI, BH, DM, MA, JB, SK, and WL provided critical revisions to the manuscript draft. All authors read and approved the final version of the manuscript draft. JYN supervised all aspects of the study.

